# Two years of longitudinal measurements of human adenovirus group F, norovirus GI and GII, rotavirus, enterovirus, enterovirus D68, hepatitis A virus, *Candida auris*, and West Nile virus nucleic-acids in wastewater solids: A retrospective study at two wastewater treatment plants

**DOI:** 10.1101/2023.08.22.23294424

**Authors:** Alexandria B. Boehm, Marlene K. Wolfe, Bradley J. White, Bridgette Hughes, Dorothea Duong

**Affiliations:** Department of Civil & Environmental Engineering, School of Engineering and Doerr School of Sustainability, Stanford University, 473 Via Ortega, Stanford, CA, USA 94305; Gangarosa Department of Environmental Health, Rollins School of Public Health, Emory University, 1518 Clifton Rd, Atlanta, GA, USA, 30322; Verily Life Sciences LLC, South San Francisco, CA, USA, 94080

## Abstract

Wastewater monitoring for infectious disease targets is increasingly used to better understand circulation of diseases. The present study validated hydrolysis-probe digital droplet (reverse-transcriptase (RT))-PCR assays for important enteric viruses (rotavirus, adenovirus group F, norovirus GI and GII, and enteroviruses), outbreak or emerging viruses (hepatitis A and West Nile virus), and an emerging drug resistant fungal pathogen (*Candida auris*). We used the assays to retrospectively measure concentrations of the targets in wastewater solids. Viral and fungal nucleic-acid concentrations were measured in two wastewater solids samples per week at two wastewater treatment plants in the San Francisco Bay Area of California, USA for 26 months. We detected all targets in wastewater solids with the exception of West Nile virus. At both wastewater treatment plants, human adenovirus group F was detected at the highest concentrations, followed by norovirus GII, enteroviruses, norovirus GI, and rotavirus at the lowest concentrations. Hepatitis A and *C. auris* were detected less consistently than the aforementioned viruses. Enterovirus D68 was detected in a limited time frame during fall 2022 at both sites. The measurements reported herein, and in some cases their seasonal trends, are consistent with previous reports of these targets in wastewater. These measurements represent some of the first quantitative measurements of these infectious disease targets in the solid fraction of wastewater. This study lays a foundation for the use of wastewater solids for the detection of specific infectious disease targets in wastewater monitoring programs aimed to better understand the spread of these diseases.

## Introduction

Infectious diseases are a leading cause of human morbidity and mortality globally^1^, and climate change is projected to exacerbate the spread of critical and emerging diseases ^2,3^. Tracking infectious disease in individuals and populations is essential to inform prevention and response, including vaccine development, vaccination campaigns, pharmaceutical and non-pharmaceutical interventions, and also to better understand disease epidemiology.

Infectious disease surveillance typically relies on clinical data. However, this data can be biased towards results from individuals with severe illness,comorbidities, and access to healthcare and diagnostic testing. In addition, clinical cases may not be reported to a centralized agency for collation, and if they are, it may take weeks for data to be organized and delivered to a public health agency for consumption. The biases and delays related to clinical data can therefore lead to missed opportunities to protect public health, particularly at critical times at the emergence of a disease, or the beginning and peak of an outbreak.

Wastewater monitoring for infectious disease surveillance can be used to assess the occurrence of diseases in populations. Wastewater contains excretions from individuals that enter drains and toilets in households and businesses including feces, urine, sputum, mucus and saliva. When individuals are infected by a pathogen, then that pathogen or its components (nucleic-acids and proteins) are excreted and often enter the wastewater. When wastewater is collected at a wastewater treatment plant (WWTP), it represents a composite sample of all the people in the sewershed that the WWTP serves (anywhere from ~10^4^ − 10^6^ individuals), including contributions of all infected individuals in the community. Unlike clinical data, this includes those who are asymptomatic or mildly symptomatic and those who do not seek or receive medical care. Results regarding the presence and concentration of different infectious disease targets (usually genomic nucleic-acids from viruses, bacteria or eukaryotes) can be available within 24 hours of sample collection for the community as a whole, limiting reporting and collating delays. Therefore, data from wastewater monitoring can be complementary to clinical data and may even provide early warnings of disease spread. Work to date suggests that wastewater monitoring can be used to detect occurrence of respiratory diseases including COVID-19^4^, influenza A^5^ and B^5,6^, respiratory syncytial virus (RSV)^7^, and seasonal coronaviruses^6^, enteric diseases including human norovirus^8^ and salmonellosis^9^, and other diseases such as mpox^10^, hepatitis A^11^, and *Candida auris*^12^ infections.

Raw wastewater is a complex mixture of liquids (water, urine, and other liquids put down drains), and solids or particles (feces, food, cells, and other material). Previous work has shown that viruses tend to partition to the solids in wastewater where their concentrations can be orders of magnitude higher than in the liquid^10,13–16^. Particles are typically defined using an operational definition (for example, larger than 0.45 µm in diameter), and using this definition, bacteria and eukaryotic organisms in wastewater are part of the solid fraction. The solid fraction can be obtained by centrifuging raw wastewater to collect a pellet, or by sampling sludge from the primary clarifier. Many wastewater monitoring programs for infectious disease targets make measurements in the liquid phase of wastewater, but a handful^17–19^, including our program, use solids where these targets are already enriched.

The goal of this study is to expand our understanding of the types of infections that may be surveilled through the use of wastewater monitoring. Herein, we conduct a retrospective study to investigate the occurrence of pathogen nucleic acids in wastewater samples over twenty-six months. Our work focuses on analyzing nucleic-acids associated with the particle/solid fraction of wastewater to take advantage of the “natural” ability of solids to concentrate the infectious disease targets of interest. In particular, we measured concentrations of human adenovirus group F, rotavirus, human norovirus GII, human norovirus GI, West Nile virus, hepatitis A virus, enteroviruses, enterovirus D 68, and *Candida auris* nucleic-acids in wastewater from two samples each week at two large wastewater treatment plants in the San Francisco Bay Area of California, USA.

## Methods

### RT-PCR assays

We designed novel (RT-)PCR primers and internal hydrolysis probes for adenovirus group F (hereafter, HAdV) and enterovirus D-68 variants circulating in the Northern hemisphere 2022 winter (EVD68-2022). In addition, we designed a novel probe for *Candida auris* to use in conjunction with previously published specific and sensitive primers^20^. To design these primers and probes, *C. auris*, HAdV, and EVD68-2022 genome sequences were downloaded from NCBI in February 2023, April 2022, and September 2022, respectively, and aligned to identify conserved regions. Primers and probes were developed *in silico* using Primer3Plus (https://primer3plus.com/). Parameters used in assay development (e.g. sequence length, GC content, and melt temperatures) are provided in Table S1. Primers and probes were then screened for specificity *in silico*, and *in vitro* against virus or fungal panels, intact viruses, synthetic viral genomic RNA, or cDNA sequences (Table 1).

**Table 1.**
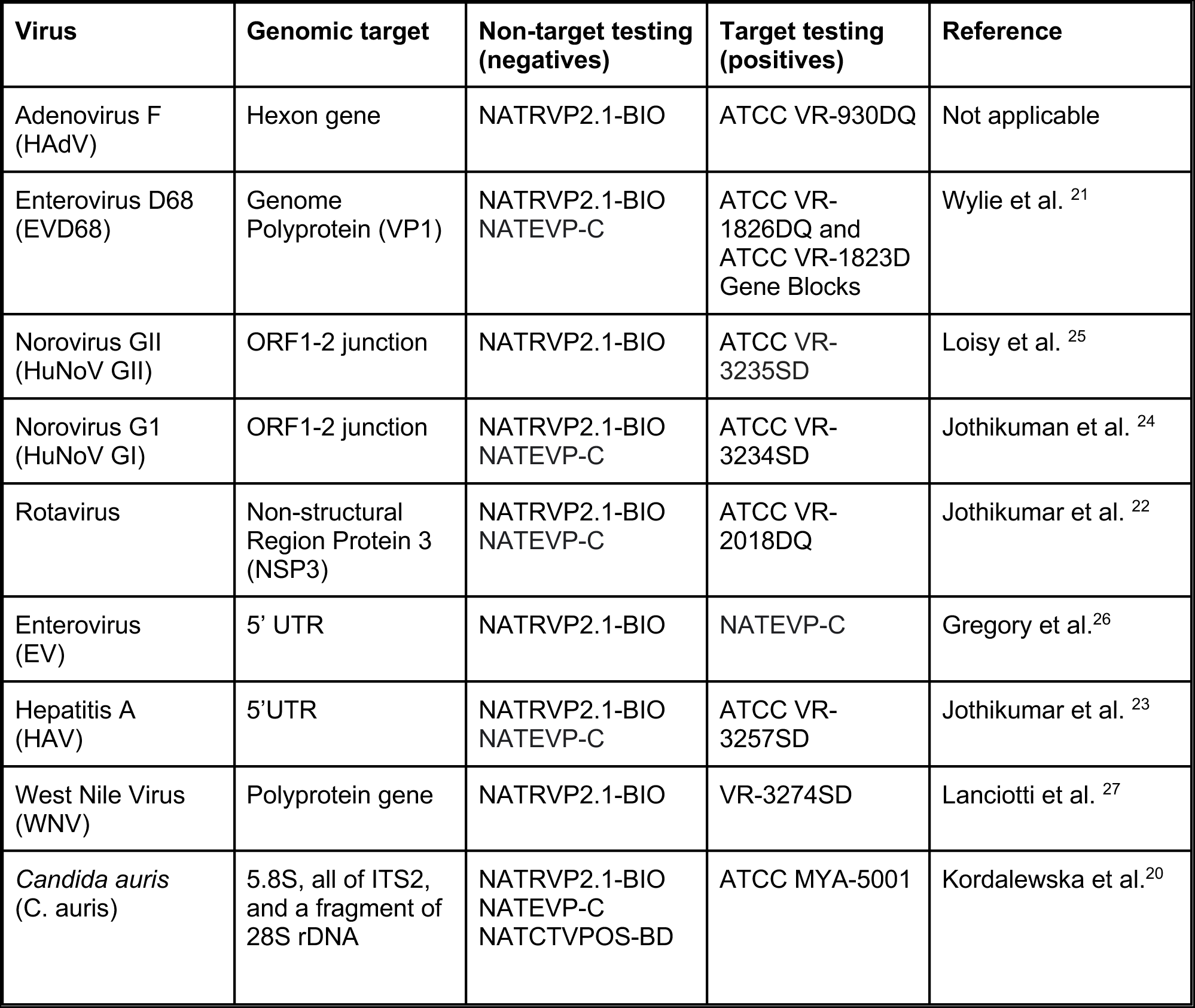
Viruses/fungus considered in this study and the region of the genome that each assay targeted. Viruses/fungi used to test specificity are indicated as “non target testing” and viruses/fungi used positive controls are indicated as “target testing”. The reference for each assay is provided aside from HAdV for which an assay was developed herein. As described in the text, we also developed novel primers and adjusted the probe for the EVD-68 assay to capture the variant circulating in the winter of 2022. All non-target controls are panels sold by Zeptomatrix (panels begin with NAT prefix, “Zepto”, Buffalo, NY). ATCC is American Type Culture Collection. The NATRVP2.1-BIO panel includes chemically inactivated intact influenza viruses, parainfluenza viruses, adenovirus, rhinovirus, metapneumovirus, and coronaviruses. The NATEVP-C panel includes chemically inactivated intact coxsackieviruses, echovirus, and parechovirus. The NATCTVPOS-BD includes chemically inactivated *Candida albicans*, *C. krusei*, *C. glabrata*, and *Trichomoniasis vaginalis*. The full list of species in the panels is available from the vendor.

| Virus | Genomic target | Non-target testing (negatives) | Target testing (positives) | Reference |
| --- | --- | --- | --- | --- |
| Adenovirus F (HAdV) | Hexon gene | NATRVP2.1-BIO | ATCC VR-930DQ | Not applicable |
| Enterovirus D68 (EVD68) | Genome Polyprotein (VP1) | NATRVP2.1-BIO<br>NATEVP-C | ATCC VR-1826DQ and<br>ATCC VR-1823D<br>Gene Blocks | Wylie et al. <sup>21</sup> |
| Norovirus GII (HuNoV GII) | ORF1-2 junction | NATRVP2.1-BIO | ATCC VR-3235SD | Loisy et al. <sup>25</sup> |
| Norovirus GI (HuNoV GI) | ORF1-2 junction | NATRVP2.1-BIO<br>NATEVP-C | ATCC VR-3234SD | Jothikuman et al. <sup>24</sup> |
| Rotavirus | Non-structural Region Protein 3 (NSP3) | NATRVP2.1-BIO<br>NATEVP-C | ATCC VR-2018DQ | Jothikumar et al. <sup>22</sup> |
| Enterovirus (EV) | 5' UTR | NATRVP2.1-BIO | NATEVP-C | Gregory et al. <sup>26</sup> |
| Hepatitis A (HAV) | 5'UTR | NATRVP2.1-BIO<br>NATEVP-C | ATCC VR-3257SD | Jothikumar et al. <sup>23</sup> |
| West Nile Virus (WNV) | Polyprotein gene | NATRVP2.1-BIO | VR-3274SD | Lanciotti et al. <sup>27</sup> |
| *Candida auris* (C. auris) | 5.8S, all of ITS2, and a fragment of 28S rDNA | NATRVP2.1-BIO<br>NATEVP-C<br>NATCTVPOS-BD | ATCC MYA-5001 | Kordalewska et al. <sup>20</sup> |

In addition to the novel assays, we used previously published assays for enterovirus D 68 (EVD68)^21^, rotavirus^22^, hepatitis A virus (HAV)^23^, human norovirus (HuNoV) GI^24^ and HuNoV GII^25^, enteroviruses (EV)^26^, and West Nile Virus (WNV)^27^. Each sample was also assayed for the SARS-CoV-2 N gene^28^, and pepper mild mottle virus M gene (PMMoV)^28^ using previously published assays.

All primers and probe sequences (including custom and previously published assays) were tested in vitro for specificity and sensitivity using virus or fungal panels (NATtrol Respiratory Verification Panel NATRVP2.1-BIO, NATtrol™ EV Panel NATRVPEVP-C, NATCTVPOS-BD), and genomic and synthetic target nucleic acids purchased from American Type Culture Collection (ATCC, Manassas, VA) or Twist Biosciences (South San Francisco, CA) (Table 1). The respiratory virus panel includes chemically inactivated intact influenza viruses, parainfluenza viruses, adenovirus, rhinovirus A, metapneumovirus, rhinovirus, RSV, several coronaviruses, and SARS-CoV-2; the EV panel includes chemically inactivated intact coxsackieviruses, echovirus, and parechovirus. The fungal panel contains chemically inactivated fungi. Nucleic acids were extracted from intact viruses or cells using Chemagic Viral DNA/RNA 300 Kit H96 for Chemagic 360 (PerkinElmer, Waltham, MA).

Nucleic acids were used undiluted as template in digital droplet (RT-)PCR singleton assays for sensitivity and specificity testing in single wells. The concentration of targets used in the in vitro specificity testing was between 10^3^ and 10^4^ copies per well. Negative (RT-)PCR controls were included on each plate.

### Wastewater analyses

Two wastewater treatment plants that serve 75% (1,500,000 people) of Santa Clara County, California (San José-Santa Clara Regional Wastewater Facility, SJ) and 25% (250,000 people) of San Francisco County (Oceanside Water Pollution Control Plant, OSP) were included in the study (Figure 1). Further descriptions of these WWTPs can be found elsewhere^10,18^.

**Figure 1.**
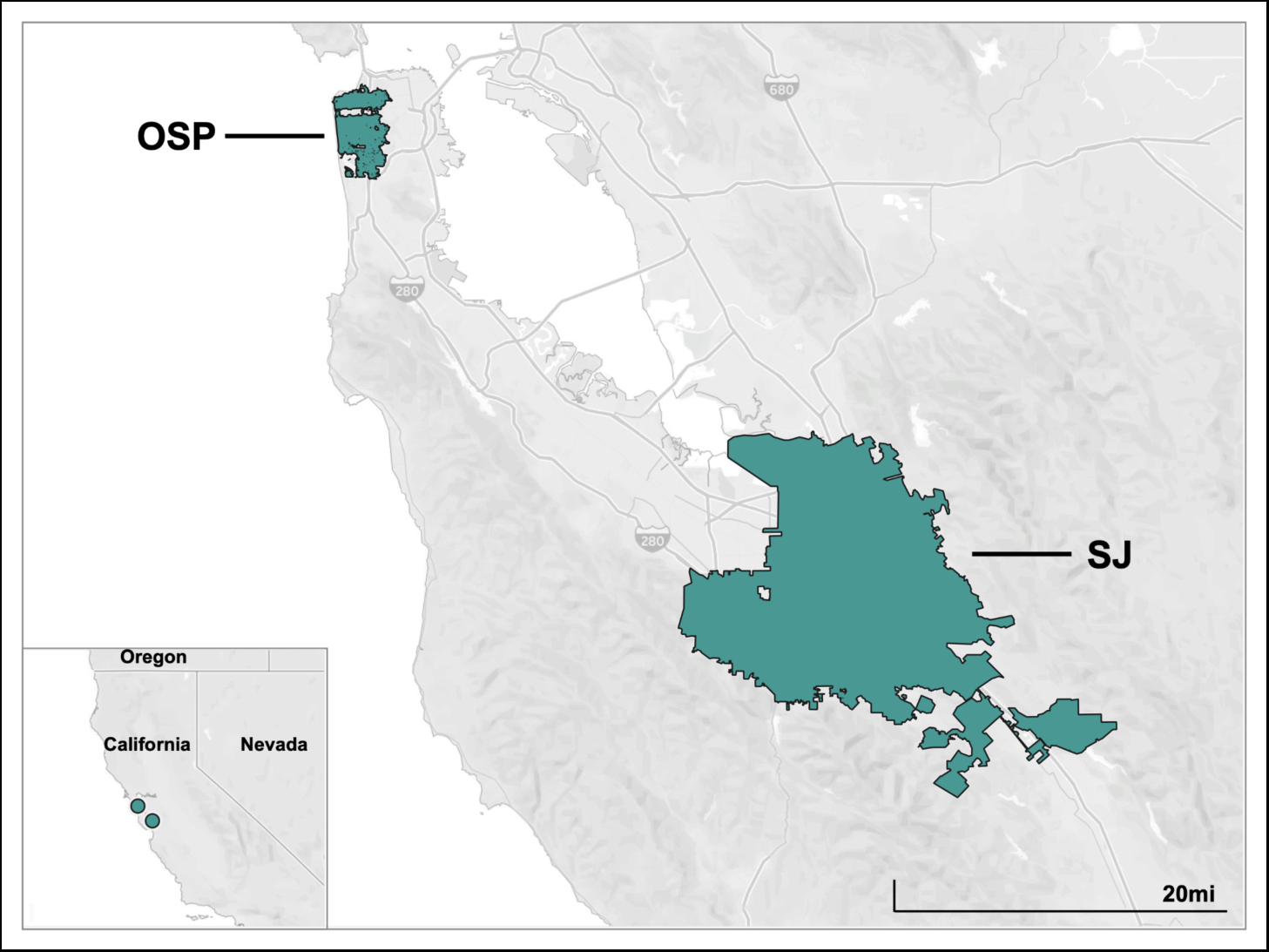
Sewersheds served by the two WWTPs in this study.

Samples were collected daily for a prospective COVID-19 wastewater monitoring effort starting in November 2020, and a subset of those samples (two samples per week, 459 samples total) are used in this study. The samples were chosen to span a 26.5 month period (2/1/21 - 4/14/23, month/day/year format) that included implementation and easing of indoor mask mandates, changes in COVID-19 vaccine availability, public health promotion of hand hygiene and mask wearing, and periods of both high and low COVID-19 incidence.

Fifty mL of settled solids were collected using sterile technique in clean bottles from the primary clarifier. At SJ, twenty-four hour composite samples were collected by combining grab samples from the sludge line every six hours. At OSP, a grab sample was collected from the sludge line each day. Samples were stored at 4°C, transported to the lab, and processed within six hours. Solids were then dewatered^29^ and frozen at −80°C for 4 - 60 weeks. Samples were thawed overnight at 4°C and then nucleic acids were obtained from the dewatered solids following previously published protocols^28,30^. Those protocols include suspending the solids in a buffer at a concentration of 75 mg/ml and using an inhibitor removal kit; together these processes alleviate potential inhibition while maintaining good assay sensitivity ^28,31^. Nucleic-acids were obtained from 10 replicate sample aliquots. Each replicate nucleic-acid extract from each sample was subsequently stored between 8 and 273 days (median 266 days) for SJ and between 1 and 8 days (median 5 days) for OSP at −80°C and subjected to a single freeze thaw cycle. Upon thawing, targets were measured immediately using using digital droplet (RT-)PCR with multiplexed assays for SARS-CoV-2, rotavirus, HAdV, HAV, HuNoV GI, HuNoV GII, WNV, EVD68, EV, *C. auris*, and PMMoV. PMMoV is highly abundant in human stool and wastewater ^32^ and is used as an internal recovery and fecal strength control^33^. Each nucleic acid extract was run in a single well so that 10 replicate wells were run for each sample for each assay.

The ddRT-PCR methods applied to wastewater solids to measure PMMoV in a singleplex reaction and are provided in detail elsewhere^18^. The human virus assays were run in multiplex using a probe-mixing approach and unique fluorescent molecules (HEX, FAM, Cy5, Cy5.5, ROX, ATTO950) in two sets of reactions. One reaction included primers and probes for rotavirus (fluorescent molecule(s) on probe: FAM), SARS-CoV-2 (FAM/HEX), WNV (ROX), HuNoV GII (ATTO590), and HAdV (ROX/ATTO590). The second included primers and probes for EVD68 (FAM), HAV (Cy5), EV (Cy5.5), HuNoV GI (ATTO590), and *C. auris* (ROX/ATTO590). Note that the two reactions contained several additional primers and probe sets that yielded results not reported herein. In particular, assays targeting the genome of HIV and several influenza subtype markers were included. Each reaction was run on its own 96-well plate.

Each 96-well PCR plate of wastewater samples included PCR positive controls for each target assayed on the plate in 1 well, PCR negative no template controls in two wells, and extraction negative controls (consisting of water and lysis buffer) in two wells. PCR positive controls consisted of viral gRNA or gene blocks (Table S1).

ddRT-PCR was performed on 20 µl samples from a 22 µl reaction volume, prepared using 5.5 µl template, mixed with 5.5 µl of One-Step RT-ddPCR Advanced Kit for Probes (Bio-Rad 1863021), 2.2 µl of 200 U/µl Reverse Transcriptase, 1.1 µl of 300 mM dithiothreitol (DDT) and primers and probes mixtures at a final concentration of 900 nM and 250 nM respectively. Primer and probes for assays were purchased from Integrated DNA Technologies (IDT, San Diego, CA) (Table 2). Human virus and fungal targets were measured in reactions with undiluted template whereas PMMoV was run on template diluted 1:100 in molecular grade water.

**Table 2.**
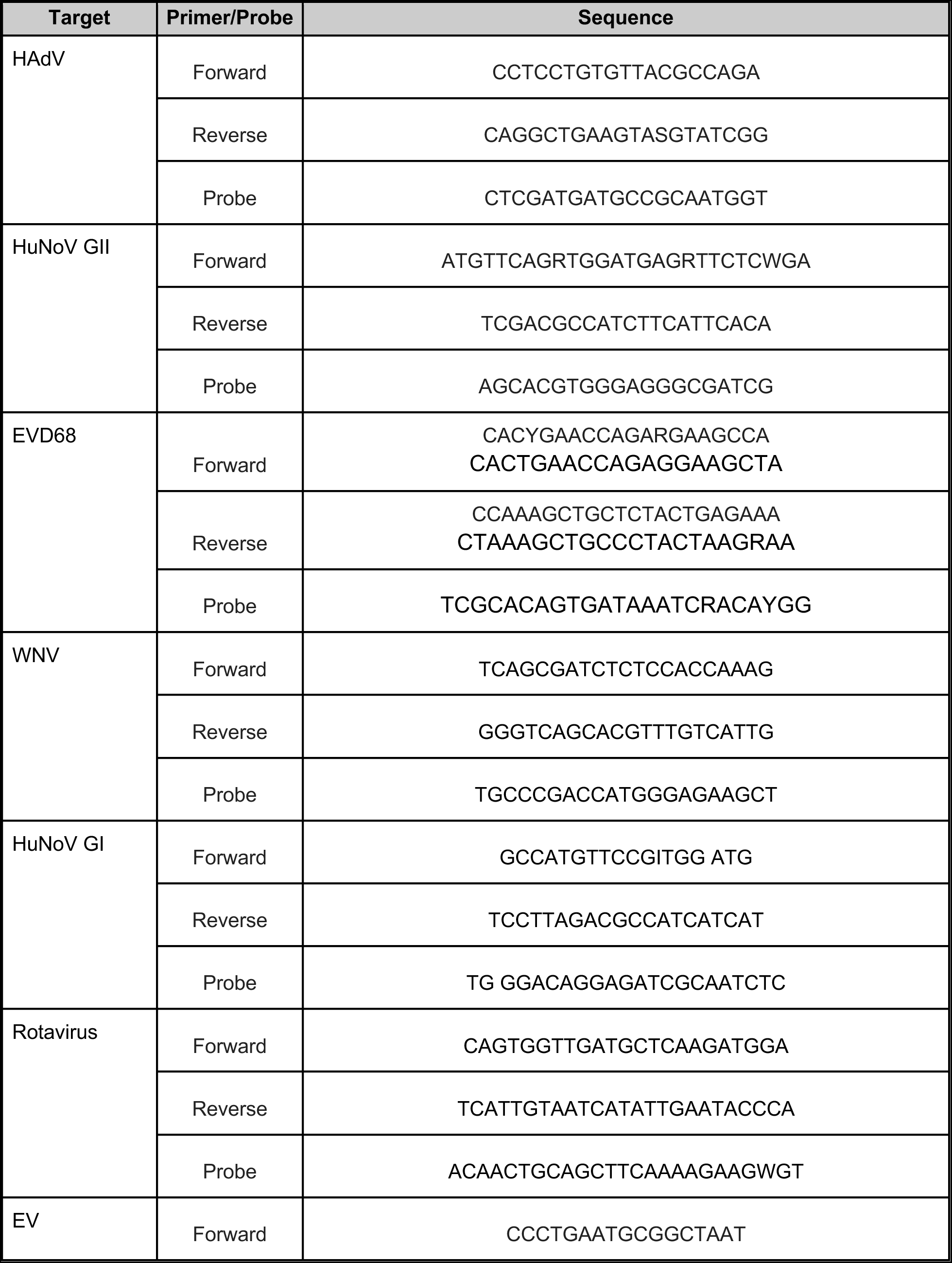
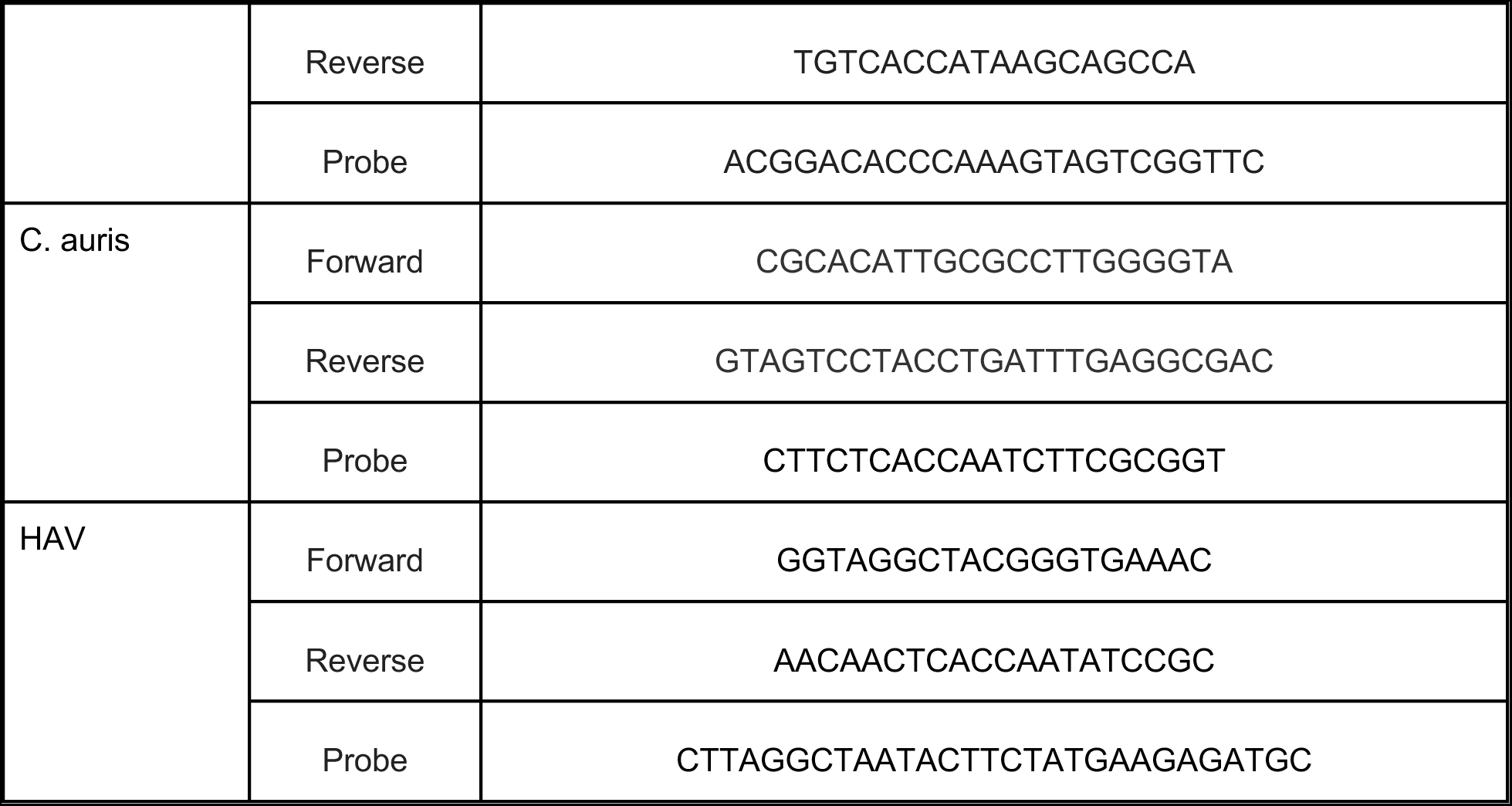
Primers and probes used in this study. Primers and probes were purchased from Integrated DNA Technologies (Coralville, IA, USA). All probes contained fluorescent molecules and quenchers (5′ HEX, FAM, Cy5, Cy5.5, ROX, and/or ATTO950/ZEN/3′ IBFQ); FAM, 6-fluorescein amidite; HEX, hexachloro-fluorescein; Cy5, Cyanine-5; Cy5.5, Cyanine5.5; ROX, carboxyrhodamine; ZEN, a proprietary internal quencher from Integrated DNA Technologies (Coralville, IA, USA); and IBFQ, Iowa Black FQ.

Droplets were generated using the AutoDG Automated Droplet Generator (Bio-Rad, Hercules, CA). PCR was performed using Mastercycler Pro (Eppendforf, Enfield, CT) with with the following cycling conditions: reverse transcription at 50°C for 60 minutes, enzyme activation at 95°C for 5 minutes, 40 cycles of denaturation at 95°C for 30 seconds and annealing and extension at 59°C (for human viruses and fungi) or 56°C (for PMMoV) for 30 seconds, enzyme deactivation at 98°C for 10 minutes then an indefinite hold at 4°C. The ramp rate for temperature changes were set to 2°C/second and the final hold at 4°C was performed for a minimum of 30 minutes to allow the droplets to stabilize. Droplets were analyzed using the QX200 or the QX600 Droplet Reader (Bio-Rad). A well had to have over 10,000 droplets for inclusion in the analysis. All liquid transfers were performed using the Agilent Bravo (Agilent Technologies, Santa Clara, CA).

Thresholding was done using QuantaSoft™ Analysis Pro Software (Bio-Rad, version 1.0.596) and QX Manager Software (Bio-Rad, version 2.0). Replicate wells were merged for analysis of each sample. In order for a sample to be recorded as positive, it had to have at least 3 positive droplets.

Concentrations of RNA targets were converted to concentrations in units of copies (cp)/g dry weight using dimensional analysis. The total error is reported as standard deviations and includes the errors associated with the Poisson distribution and the variability among the 10 replicates. Three positive droplets across 10 merged wells corresponds to a concentration between ~500-1000 cp/g; the range in values is a result of the range in the equivalent mass of dry solids added to the wells. Data collected as part of the retrospective study are available from the Stanford Digital Repository (https://purl.stanford.edu/qt551tn4819).

### Multiplex assay performance

Digital droplet PCR is well suited for multiplexing. This is because of the inherent design of the method; there is unlikely to be more than one target nucleic acid per droplet when assaying samples for rare targets like human pathogens. Regardless, we sought to confirm that the multiplexing up to eight assays in one well did not interfere with the quantification of targets. In particular, we tested whether the quantification of a single target in the presence of and absence of relatively high concentrations of the 7 other targets was substantially different. To do so, we first quantified three decimal dilutions of a target nucleic acid in the absence of any other targets using the (RT)-PCR chemistry that included primers and probes for all 8 targets. Then, we quantified the same decimal dilutions of the single target in the presence of relatively high concentrations of standards of the 7 other targets. Each dilution was run in a single well and no template, negative controls were included on each PCR plate. Assays were run and thresholded as described above. Results were expressed as copies per reaction and the 68% confidence intervals, as output by the instrument, were included.

### SARS-CoV-2 and PMMoV RNA concentrations from prospective study in fresh samples

In order to understand potential losses from storage and freeze thaw of the samples and their nucleic acid extracts, we compared measurements of the SARS-CoV-2 N gene and PMMoV RNA measured in this study obtained using the stored samples to measurements from the same samples that were not stored and analyzed prospectively for a large monitoring effort; samples were analyzed within hours of sample collection. The methods for analysis for the N gene through 3/13/23 were identical to those previously described in a Data Descriptor^28^ with the exact same approaches described in this paper except the assay was multiplexed with two other assays using a two color droplet reader (QX200, Bio-rad). Between 3/13/23 and 4/15/23, the N gene was multiplexed with assays for five adjacent SNPs in SARS-CoV-2 XBB*, respiratory syncytial virus (RSV), influenza A, influenza B, and HuNoV GII in conjunction with a 6-color droplet reader (QX600, Bio-rad).

## Results & Discussion

### QA/QC

Results are reported as suggested in the Environmental Microbiology Minimal Information guidelines^34^ (Figure S2). Extraction and PCR negative and positive controls performed as expected (negative and positive, respectively). PMMoV measurements were used to assess potential for gross extraction failures as it is present in very high concentrations in the samples naturally, and lack of its detection, or abnormally low measurements might indicate gross extraction failures. The median (interquartile range) log_10_-transformed PMMoV was 9.1 (9.0-9.2) and 8.7 (8.6-8.8) log_10_ copies/g at SJ and OSP, respectively. The lowest measurements at the two sites were 8.5 (SJ) and 8.1 (OSP) log_10_ copies/g and given these lowest values are within an order of magnitude of the medians, we concluded that there was no gross extraction failures. We opted to not use an exogenous viral control, like bovine coronavirus, owing to the complexities associated with interpreting recovery of an exogenous spiked control in environmental samples^35^, and its potential to interfere with other uses of the samples, for example viral metagenomics.

We compared measurements of the N gene and PMMoV made on the samples that were fresh to those used in this study that were stored, and for which the RNA underwent one freeze thaw. Median ratio of PMMoV measurements made in this study to those made using fresh samples was 0.9 (0.6-1.2) at SJ and 1.0 (0.7-1.4) at OSP suggesting limited degradation of the PMMoV target in the stored and freeze thawed samples. Median ratio of SARS-CoV-2 N gene measurements made in this study to those made using fresh samples was 0.2 (0.1-0.3) at SJ and 0.4 (0.2-0.6) at OSP suggesting storage and freeze thaw may have reduced measurement concentrations, but by less than an order of magnitude. Although not ideal, storage of samples is essential for retrospective work like this.

### Assay sensitivity and specificity

We tested previously published assays for sensitivity and specificity. In silico analysis indicated no cross reactivity of the (RT-)PCR probe-based assays (Table 2) with sequences deposited in National Center for Biotechnology Information (NCBI); in vitro testing against non-target and target viral or fungal nucleic-acids also indicated no cross reactivity. The exception was the previously published assay for EVD68 developed by Wylie et al.^21^ in 2015. In silico and in vitro testing indicated the assay was not able to detect the variant of EVD68 circulating in winter 2022 (EVD68-2022). We therefore developed a new set of primers, and modified the probe to include a degeneracy, to amplify and detect the same region of the VP1 gene targeted by the Wylie et al. assay. The EVD68 assay used herein therefore includes two forward and reverse primers in order to ensure detection of all EVD68 variants in NCBI including EVD68-2022. We confirmed that the new assay, with two sets of probes and the modified probe, was sensitive and specific in silico and in vitro to both the EVD68-2022 strains and the EVD68 historical strains.

We developed a new assay for HAdV. We confirmed assay sensitivity and specificity in silico. In vitro, there was no cross reactivity between the HAdV assay and non-target viral genomic nucleic acids. We developed a probe to use in conjunction with previously published primers for *C. auris*. The newly designed assay was also sensitivity and specific based on in vitro and in silico analysis.

In this project, we multiplexed eight different hydrolysis-probe assays in each reaction. We tested whether quantification of a target in the multiplex was affected by the presence of the 7 other nucleic-acid targets. We found that the concentration of each target was not affected by the presence of a high concentration or orthogonal targets as background (Figure 2). This is evidenced by the fact that target nucleic-acid concentrations measured in the absence and presence of background orthogonal nucleic acids are similar across the decimal dilutions tested.

**Figure 2.**
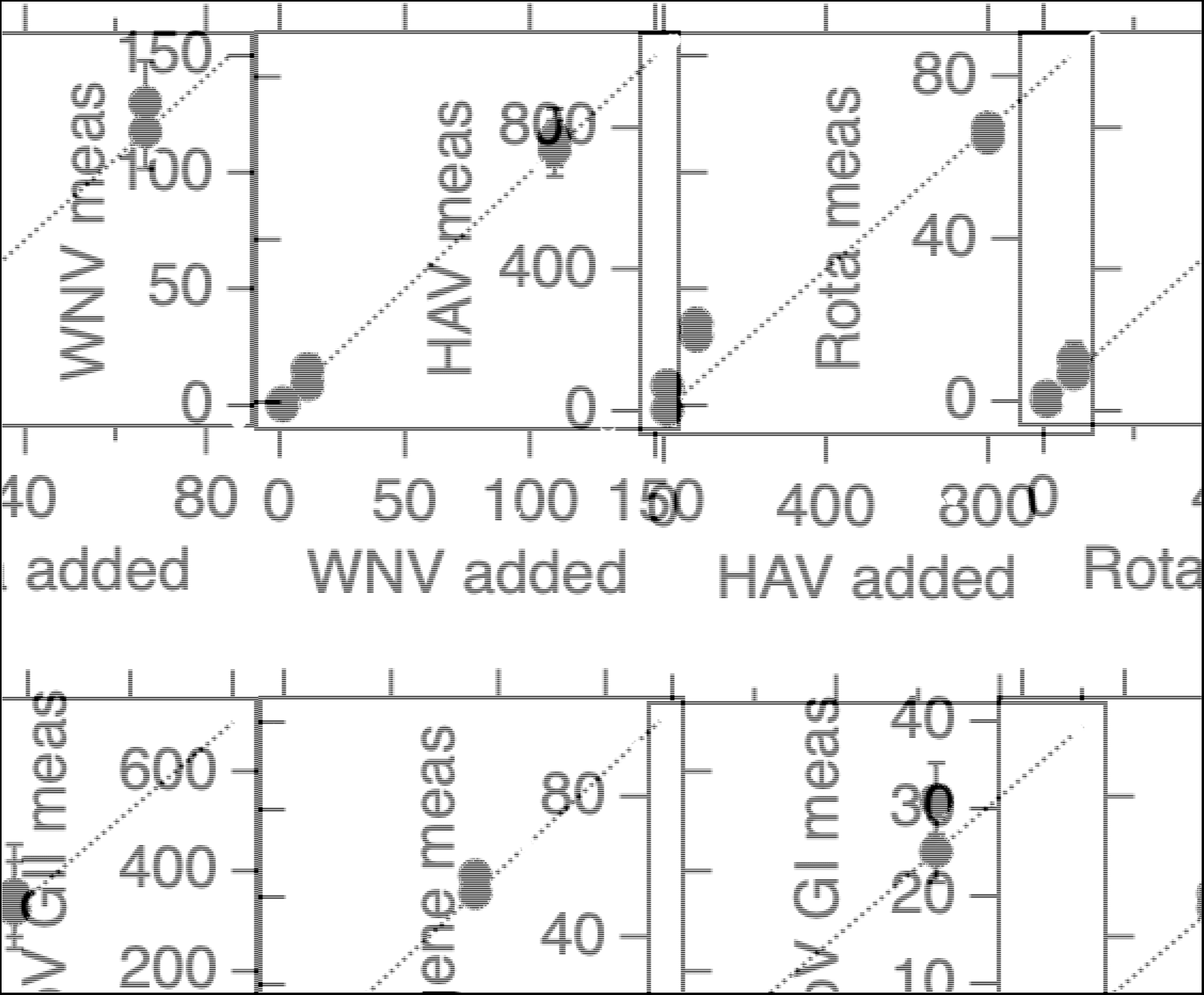
Multiplex assay performance. The concentrations of the different targets in units of copies per reaction input and measured in each experiment is provided. The white symbols are for reactions without the 7 background nucleic-acid targets and black symbols include high concentrations of the 7 other nucleic-acid targets. Error bars are standard deviations, if error bars cannot be seen, then they are smaller than the symbol. The line represents the 1:1 line. “Meas” is measured, “Rota” is rotavirus. Abbreviations for the different targets are provided in the main text.

### Enteric viruses in wastewater solids

Concentrations of EV, HAdV, rotavirus, HuNoV GI, and HuNoV GII nucleic-acids in wastewater solids over the project are shown in Figure 3. Figure S2 shows concentrations normalized by PMMoV. All targets were detected in wastewater solids throughout the study with most showing season patterns with higher concentrations in the winter 2021-2022 and 2022-2023, but not in winter 2020-2021. The exception is EV which was present even at relatively high concentrations in winter 2020-2021. Viral enteric infections are seasonal with higher rates in the winter months^36^, so the seasonal patterns in the wastewater are consistent with that. The lack of a winter peak in the winter of 2020-2021 may be a result of the non-pharmaceutical interventions in place during the COVID-19 pandemic which may have limited the spread of different infections.

**Figure 3.**
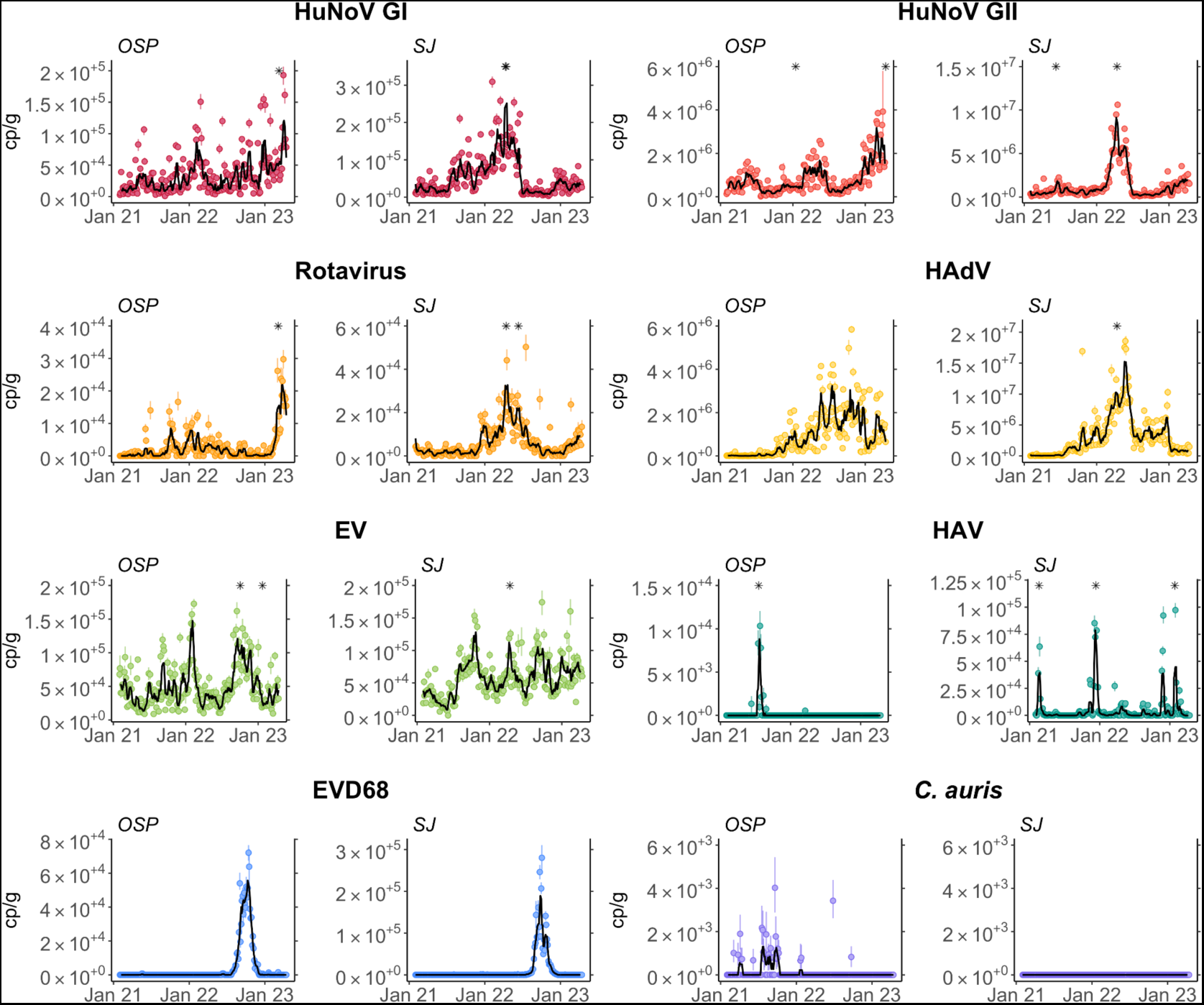
Concentrations of HuNoV G1 and GII, rotavirus, HAdV, EV, HAV, EVD68 and *C. auris* nucleic-acids in wastewater solids at OSP and SJ. Error bars show standard deviation. The black line shows a 5-adjacent sample trimmed average. A gray symbol at the top of the plot indicates a value that is higher than the y-axis scale.

Median concentrations of each enteric viral target were significantly higher at SJ relative to OSP (Mann-Whitney U Test, all p<0.05) but the effect size was less than 1 order of magnitude (Table 3). Interestingly, median concentrations of PMMoV were also higher at SJ, and the normalized concentrations of these targets are more comparable (Figure S2). Within each WWTP, median concentrations of the different viruses ranked highest to lowest were HAdV (highest median concentration of 5.8×10^5^ and 1.9×10^6^ cp/g for OSP and SJ, respectively) followed by HuNoV GII (5.7×10^5^ and 7.0×10^5^ cp/g for OSP and SJ, respectively), EV (4.1×10^4^ and 5.4×10^4^ cp/g for OSP and SJ, respectively), HuNoV GI (2.8×10^4^ and 3.4×10^4^ cp/g for OSP and SJ, respectively), and rotavirus with the lowest (1×10^3^ and 3.3 x 10^3^ cp/g for OSP and SJ, respectively) (Table 3). Other summary statistics (25th and 75th percentiles, maximums and number of non-detects) generally follow the same pattern.

**Table 3.** Summary statistics for infectious disease targets in OSP and SJ wastewater solids. Units of concentration are copies per gram dry weight. 25th is 25th percentile, 75th is 75th percentile, max is the maximum value, # ND is number of non-detect, # is the total number of samples.

| WWTP |  | Rota | HAdV | EV | HuNoV GI | HuNoV GII | HAV | EVD68 | C. auris |
| --- | --- | --- | --- | --- | --- | --- | --- | --- | --- |
| OSP | median | 1.0x10<sup>3</sup> | 5.8x10<sup>5</sup> | 4.1x10<sup>4</sup> | 2.8x10<sup>4</sup> | 5.7x10<sup>5</sup> | 0 | 0 | 0 |
|  | 25th | 0 | 4.9x10<sup>4</sup> | 2.6x10<sup>4</sup> | 1.2x10<sup>4</sup> | 3.5x10<sup>5</sup> | 0 | 0 | 0 |
|  | 75th | 3.2x10<sup>3</sup> | 1.5x10<sup>6</sup> | 6.9x10<sup>4</sup> | 4.7x10<sup>4</sup> | 1.1x10<sup>6</sup> | 0 | 0 | 0 |
|  | max | 7.8x10<sup>4</sup> | 5.8x10<sup>6</sup> | 2.4x10<sup>5</sup> | 2.6x10<sup>5</sup> | 6.4x10<sup>6</sup> | 9x10<sup>4</sup> | 7.2x10<sup>4</sup> | 4.0x10<sup>3</sup> |
|  | # ND | 89 | 9 | 0 | 0 | 0 | 220 | 182 | 202 |
|  | # | 230 | 230 | 230 | 230 | 230 | 230 | 230 | 230 |
| SJ | median | 3.3x10<sup>3</sup> | 1.9x10<sup>6</sup> | 5.4x10<sup>4</sup> | 3.4x10<sup>4</sup> | 7.0x10<sup>5</sup> | 6.6x10<sup>2</sup> | 0 | 0 |
|  | 25th | 1.5x10<sup>3</sup> | 5.3x10<sup>5</sup> | 3.7x10<sup>4</sup> | 1.6x10<sup>4</sup> | 3.5x10<sup>5</sup> | 0 | 0 | 0 |
|  | 75th | 8.5x10<sup>3</sup> | 4.3x10<sup>6</sup> | 7.5x10<sup>4</sup> | 7.6x10<sup>4</sup> | 1.5x10<sup>6</sup> | 3.1x10<sup>3</sup> | 0 | 0 |
|  | max | 2.7x10<sup>5</sup> | 8.5x10<sup>7</sup> | 1.2x10<sup>6</sup> | 1.4x10<sup>6</sup> | 1.1x10<sup>8</sup> | 4.0x10<sup>5</sup> | 2.8x10<sup>5</sup> | 0 |
|  | # ND | 24 | 0 | 1 | 0 | 0 | 111 | 177 | 229 |
|  | # | 229 | 229 | 229 | 229 | 229 | 229 | 229 | 229 |

Rotavirus has previously been reported to be present in wastewater sludge around the world^37–39^, but quantitative data on its concentration, to our knowledge, have not been reported. In the present study, we observed concentrations in wastewater solids up to 10^5^ cp/g with median concentrations on the order of 10^3^ cp/g. Concentrations of rotavirus, using both molecular methods and microscopy, in raw sewage has been previously reported to be between 1 and 10^5^ cp/ml^40–42^ and as high as 100 fluorescent foci per ml^43,44^ with some authors reporting seasonal occurrence^44^. The measurements in this study also exhibited a seasonal occurrence with higher concentrations in the winter-spring season. A number of studies have investigated different genotypes of rotavirus in sewage using sequencing methods^41^ and this may be an interesting area for further research within wastewater-based epidemiology applications. In our study area, oral, live rotavirus vaccines are administered to infants less than 32 weeks of age, and as a result, cases of rotavirus have decreased since vaccine introduction^45^. Based on available sequences of the rotavirus vaccine, the rotavirus assay used in our study may amplify and detect genomic RNA from the vaccine. As infants in our study area wear diapers, including both disposable and cloth, it is unclear to what extent the vaccine strains from infants will appear in wastewater and contribute to the measurements reported herein. However, as vaccine strains are live, transmission to others who do use the wastewater system may be possible^46^, with their shedding possibly influencing concentrations of rotavirus RNA in the waste stream.

As with rotavirus, there are limited reports of HuNoV GII and GI in wastewater solids^37^ with some exceptions. Boehm et al.^16^ report concentrations of HuNoV GII in wastewater solids from these same plants from a different set of samples and found similar concentrations, but they did not report concentrations of HuNoV GI. There is a large number of studies reporting HuNoV in raw sewage, many of which are described in a systematic review by Eftim et al.^47^ They report median concentrations globally of 10-100 cp/ml with concentrations of HuNoV GII higher than GI, particularly in North America. This is consistent with our findings that indicate that HuNoV GI is present at an order of magnitude lower concentrations than HuNoV GII. Seasonal variation of HuNoV in sewage has been reported^8,47^, also consistent with the higher levels we observe in wastewater solids in this study in the winter to spring.

The EV assay we used in this study is a pan-Enterovirus genus assay and therefore detects a number of different viruses including coxsackieviruses, enteroviruses, and echoviruses. The diversity of viruses detected with this assay, and the diversity of diseases associated with them, including meningitis, diarrheal disease, hand-foot-and-mouth disease, and respiratory disease, may explain why the concentrations observed in the present study lack clear seasonal patterns as might be expected for viruses that cause single diseases^36^. Enteroviruses have been measured extensively in wastewater^48,49^, and their presence in sludge has been documented^13,50^. Monpoeho et al.^50^ reported up to 23,000 copies of EV RNA per gram of sludge, strikingly consistent with the concentrations observed in this study (median ~ 10^4^ cp/g). Concentrations of enteroviruses in raw liquid sewage can range from non-detect to 1000 cp/ml depending on location^48^. Enteroviruses have been shown to preferentially adsorb to wastewater solids^13^, but the degree of sorption is governed by the type of enterovirus^51^.

The HAdV assay used in this study targets group F which includes adenovirus 40 and 41; both cause diarrheal illness. HAdV has been documented at relatively high concentrations in raw sewage (~10^3^ viruses/ml) at locations around the world^52–54^. Consistent with those reports, we also found HAdV at the highest concentration of all the viruses tested in wastewater solids in this study. HAdV has previously been reported to preferentially adsorb to wastewater solids^13^ where it can be found at concentrations 10^4^ higher than in the liquid phase on a per mass basis. Martin et al.^55^ and Reyne et al.^56^ reported that concentrations of HAdV F41 in wastewater influent in Ireland correlated with laboratory confirmed adenovirus F41 infections and showed a seasonal trend in both, with high occurrence of infections and high concentrations in wastewater in the winter-spring seasons. We also observed some evidence of seasonal changes in HAdV concentrations in wastewater solids, but that pattern appears to be more pronounced in SJ compared to OSP.

### Hepatitis A virus (HAV)

We detected HAV in approximately 50% of the wastewater solids samples from SJ, but only 5% of the samples from OSP. Median concentrations were non-detect at OSP and 6.6 x 10^2^ cp/g at SJ with little evidence of seasonality. The lack of clear seasonality is consistent with reports that HAV infections typically are not seasonal^57^. HAV has been detected in raw sewage globally^58^. Hellmer et al ^59^ measured HAV in wastewater influent up to 14 cp/ml ^59^, and found genotypically identical strains in wastewater and clinical samples. Like our measurements, they found a high degree of variability in their measurements and suggested that due to high levels of shedding of the virus by infected patients, even a single infection could cause large spikes in wastewater concentrations followed by non-detectable concentrations. McCall et al.^11^ measured HAV in wastewater influent during a HAV outbreak in Michigan and report median concentrations of 10^3^ cp/ml; the authors also showed a significant positive correlation between wastewater measurements and the number of laboratory confirmed incident HAV cases. There is little data on HAV in wastewater solids; in their review, Yin et al.^13^ identified one study^60^ that indicated HAV partitioning to solids.

### Enterovirus D68 (EVD68)

EVD68 was non-detect in all samples except for those collected in the fall 2022. During this season, EVD68 RNA concentrations increased coherently and dramatically across the two WWTPs over ~ 6 weeks starting in mid-August to reach a peak in early October 2022, and then reaching non-detect again by the end of December 2022. The highest concentration observed was 72,000 cp/g at OSP and 280,000 cp/g at SJ.

A similar seasonal pattern in EVD68 in wastewater in 2022 was reported in Texas^61^, as well as in Israel^62^ and the United Kingdom^63^. The study in Israel reports good agreement between wastewater concentrations of EVD68 and confirmed laboratory cases of EVD68 infections^62^. These previous studies did not report EVD68 nucleic acid concentrations in externally valid units (they reported presence/absence or copies per reaction, or proportions of all viruses), so a comparison of concentrations to those reported herein is not possible. EVD68 adsorption to wastewater solids has not been previously reported.

### Candida auris

*C. auris* was not detected in any sample from SJ, but was detected in 28 of 230 (12%) samples from OSP. The highest concentration detected at OSP was 4000 cp/g. Detection of *C. auris* at OSP was most frequent between mid July 2021 and mid October 2021. *C. auris* is a drug resistant fungal pathogen that emerged in 2009^64^. Previous studies have reported detection of *C. auris* in wastewater influent at concentrations of 1 to 1000 cp/ml, and suggested its presence is associated with confirmed infections and illnesses in the communities contributing to the wastewater^12^. *C. auris* is 2.5 - 5.0 µm in diameter^65^ and thus is, by definition, a particle and is expected to be in the solid fraction of wastewater.

### West Nile Virus (WNV)

WNV is an arbovirus spread by mosquitoes. We did not detect WNV nucleic-acids in any samples - all measurements were below the lower limit of detection. This finding is consistent with the lack of reporting of WNV infections in the communities served by the two WWTPs (https://westnile.ca.gov/). To our knowledge, WNV nucleic acids have not been detected in wastewater. Lee et al.^66^ recently reported on challenges and opportunities associated with arbovirus surveillance using wastewater.

## Conclusion

This study validated and implemented a suite of digital (RT-)PCR assays to detect human rotavirus, adenovirus group F, norovirus GI and GII, West Nile virus, hepatitis A virus, enterovirus, enterovirus D68, and the fungal pathogen C. auris. The work presented here builds upon previous research illustrating these targets are present in wastewater, and lays a foundation for the use of solids for the detection of these infectious disease targets.

## Data Availability

https://purl.stanford.edu/qt551tn4819

## Acknowledgements

We acknowledge the numerous people at San Jose and Oceanside wastewater treatment plants who contributed to wastewater sample collection and Allegra Koch for her support with literature review. This research was performed on the ancestral and unceded lands of the Muwekma Ohlone people. We pay our respects to them and their Elders, past and present, and are grateful for the opportunity to live and work here.

## References

(1) Bhutta, Z. A.; Sommerfeld, J.; Lassi, Z. S.; Salam, R. A.; Das, J. K. Global Burden, Distribution, and Interventions for Infectious Diseases of Poverty. Infectious Diseases of Poverty 2014, 3 (1), 21. 10.1186/2049-9957-3-21.

(2) Lafferty, K. D. The Ecology of Climate Change and Infectious Diseases. Ecology 2009, 90 (4), 888–900. 10.1890/08-0079.1.

(3) Altizer, S.; Ostfeld, R. S.; Johnson, P. T. J.; Kutz, S.; Harvell, C. D. Climate Change and Infectious Diseases: From Evidence to a Predictive Framework. Science 2013, 341 (6145), 514–519. 10.1126/science.1239401.

(4) Wu, F.; Zhang, J.; Xiao, A.; Gu, X.; Lee, W. L.; Armas, F.; Kauffman, K.; Hanage, W.; Matus, M.; Ghaeli, N.; Endo, N.; Duvallet, C.; Poyet, M.; Moniz, K.; Washburne, A. D.; Erickson, T. B.; Chai, P. R.; Thompson, J.; Alm, E. J. SARS-CoV-2 Titers in Wastewater Are Higher than Expected from Clinically Confirmed Cases. mSystems 2020, 5 (4), e00614–20. 10.1128/mSystems.00614-20.

(5) Mercier, E.; D’Aoust, P. M.; Thakali, O.; Hegazy, N.; Jia, J.-J.; Zhang, Z.; Eid, W.; Plaza-Diaz, J.; Kabir, M. P.; Fang, W.; Cowan, A.; Stephenson, S. E.; Pisharody, L.; MacKenzie, A. E.; Graber, T. E.; Wan, S.; Delatolla, R. Municipal and Neighbourhood Level Wastewater Surveillance and Subtyping of an Influenza Virus Outbreak. Scientific Reports 2022, 12 (1), 15777. 10.1038/s41598-022-20076-z.

(6) Boehm, A. B.; Hughes, B.; Duong, D.; Chan-Herur, V.; Buchman, A.; Wolfe, M. K.; White, B. J. Wastewater Concentrations of Human Influenza, Metapneumovirus, Parainfluenza, Respiratory Syncytial Virus, Rhinovirus, and Seasonal Coronavirus Nucleic-Acids during the COVID-19 Pandemic: A Surveillance Study. The Lancet Microbe 2023, 4 (5), e340–e348. 10.1016/S2666-5247(22)00386-X.

(7) Hughes, B.; Duong, D.; White, B. J.; Wigginton, K. R.; Chan, E. M. G.; Wolfe, M. K.; Boehm, A. B. Respiratory Syncytial Virus (RSV) RNA in Wastewater Settled Solids Reflects RSV Clinical Positivity Rates. Environ. Sci. Technol. Lett. 2022, 9 (2), 173–178. 10.1021/acs.estlett.1c00963.

(8) Guo, Y.; Li, J.; O’Brien, J.; Sivakumar, M.; Jiang, G. Back-Estimation of Norovirus Infections through Wastewater-Based Epidemiology: A Systematic Review and Parameter Sensitivity. Water Research 2022, 219, 118610. 10.1016/j.watres.2022.118610.

(9) Diemert, S.; Yan, T. Clinically Unreported Salmonellosis Outbreak Detected via Comparative Genomic Analysis of Municipal Wastewater Salmonella Isolates. Appl. Environ. Microbiol. 2019, 85 (10), e00139–19. 10.1128/AEM.00139-19.

(10) Wolfe, M. K.; Yu, A. T.; Duong, D.; Rane, M. S.; Hughes, B.; Chan-Herur, V.; Donnelly, M.; Chai, S.; White, B. J.; Vugia, D. J.; Boehm, A. B. Use of Wastewater for Mpox Outbreak Surveillance in California. N Engl J Med 2023. 10.1056/NEJMc2213882.

(11) McCall, C.; Wu, H.; O’Brien, E.; Xagoraraki, I. Assessment of Enteric Viruses during a Hepatitis Outbreak in Detroit MI Using Wastewater Surveillance and Metagenomic Analysis. Journal of Applied Microbiology 2021, 131 (3), 1539–1554. 10.1111/jam.15027.

(12) Barber, C.; Crank, K.; Papp, K.; Innes, G. K.; Schmitz, B. W.; Chavez, J.; Rossi, A.; Gerrity, D. Community-Scale Wastewater Surveillance of Candida Auris during an Ongoing Outbreak in Southern Nevada. Environ. Sci. Technol. 2023, 57 (4), 1755–1763. 10.1021/acs.est.2c07763.

(13) Yin, Z.; Voice, T.; Tarabara, V.; Xagoraraki, I. Sorption of Human Adenovirus to Wastewater Solids. Journal of Environmental Engineering 2018, 144 (11), 06018008. 10.1061/(ASCE)EE.1943-7870.0001463.

(14) Wolfe, M. K.; Duong, D.; Bakker, K. M.; Ammerman, M.; Mortenson, L.; Hughes, B.; Arts, P.; Lauring, A. S.; Fitzsimmons, W. J.; Bendall, E.; Hwang, C. E.; Martin, E. T.; White, B. J.; Boehm, A. B.; Wigginton, K. R. Wastewater-Based Detection of Two Influenza Outbreaks. Environ. Sci. Technol. Lett. 2022, 9 (8), 687–692. 10.1021/acs.estlett.2c00350.

(15) Kim, S.; Kennedy, L. C.; Wolfe, M. K.; Criddle, C. S.; Duong, D. H.; Topol, A.; White, B. J.; Kantor, R. S.; Nelson, K. L.; Steele, J. A.; Langlois, K.; Griffith, J. F.; Zimmer-Faust, A. G.; McLellan, S. L.; Schussman, M. K.; Ammerman, M.; Wigginton, K. R.; Bakker, K. M.; Boehm, A. B. SARS-CoV-2 RNA Is Enriched by Orders of Magnitude in Primary Settled Solids Relative to Liquid Wastewater at Publicly Owned Treatment Works. Environ. Sci.: Water Res. Technol. 2022. 10.1039/D1EW00826A.

(16) Boehm, A. B.; Wolfe, M. K.; White, B.; Hughes, B.; Duong, D.; Banaei, N.; Bidwell, A. Human Norovirus (HuNoV) GII RNA in Wastewater Solids at 145 United States Wastewater Treatment Plants: Comparison to Positivity Rates of Clinical Specimens and Modeled Estimates of HuNoV GII Shedders. Journal of Exposure Science & Environmental Epidemiology 2023. 10.1038/s41370-023-00592-4.

(17) D’Aoust, P. M.; Mercier, E.; Montpetit, D.; Jia, J.-J.; Alexandrov, I.; Neault, N.; Baig, A. T.; Mayne, J.; Zhang, X.; Alain, T.; Langlois, M.-A.; Servos, M. R.; MacKenzie, M.; Figeys, D.; MacKenzie, A. E.; Graber, T. E.; Delatolla, R. Quantitative Analysis of SARS-CoV-2 RNA from Wastewater Solids in Communities with Low COVID-19 Incidence and Prevalence. Water Research 2021, 188, 116560. 10.1016/j.watres.2020.116560.

(18) Wolfe, M. K.; Topol, A.; Knudson, A.; Simpson, A.; White, B.; Duc, V.; Yu, A.; Li, L.; Balliet, M.; Stoddard, P.; Han, G.; Wigginton, K. R.; Boehm, A. High-Frequency, High-Throughput Quantification of SARS-CoV-2 RNA in Wastewater Settled Solids at Eight Publicly Owned Treatment Works in Northern California Shows Strong Association with COVID-19 Incidence. mSystems 2021, 6 (5), e00829–21. 10.1128/mSystems.00829-21.

(19) Peccia, J.; Zulli, A.; Brackney, D. E.; Grubaugh, N. D.; Kaplan, E. H.; Casanovas-Massana, A.; Ko, A. I.; Malik, A. A.; Wang, D.; Wang, M.; Warren, J. L.; Weinberger, D. M.; Omer, S. B. SARS-CoV-2 RNA Concentrations in Primary Municipal Sewage Sludge as a Leading Indicator of COVID-19 Outbreak Dynamics. Nature Biotechnology 2020, 38, 1164–1167.

(20) Kordalewska, M.; Zhao, Y.; Lockhart, S. R.; Chowdhary, A.; Berrio, I.; Perlin, D. S. Rapid and Accurate Molecular Identification of the Emerging Multidrug-Resistant Pathogen Candida Auris. Journal of Clinical Microbiology 2017, 55 (8), 2445–2452. 10.1128/jcm.00630-17.

(21) Wylie, T. N.; Wylie, K. M.; Buller, R. S.; Cannella, M.; Storch, G. A. Development and Evaluation of an Enterovirus D68 Real-Time Reverse Transcriptase PCR Assay. Journal of Clinical Microbiology 2015, 53 (8), 2641–2647. 10.1128/jcm.00923-15.

(22) Jothikumar, N.; Kang, G.; Hill, V. R. Broadly Reactive TaqMan® Assay for Real-Time RT-PCR Detection of Rotavirus in Clinical and Environmental Samples. Journal of Virological Methods 2009, 155 (2), 126–131. 10.1016/j.jviromet.2008.09.025.

(23) Jothikumar, N.; Cromeans, T. L.; Sobsey, M. D.; Robertson, B. H. Development and Evaluation of a Broadly Reactive TaqMan Assay for Rapid Detection of Hepatitis A Virus. Applied and Environmental Microbiology 2005, 71 (6), 3359–3363. 10.1128/AEM.71.6.3359-3363.2005.

(24) Jothikumar Narayanan; Lowther James A.; Henshilwood Kathleen; Lees David N.; Hill Vincent R.; Vinjé Jan. Rapid and Sensitive Detection of Noroviruses by Using TaqMan-Based One-Step Reverse Transcription-PCR Assays and Application to Naturally Contaminated Shellfish Samples. Applied and Environmental Microbiology 2005, 71 (4), 1870–1875. 10.1128/AEM.71.4.1870-1875.2005.

(25) Loisy, F.; Atmar, R. L.; Guillon, P.; Le Cann, P.; Pommepuy, M.; Le Guyader, F. Real-Time RT-PCR for Norovirus Screening in Shellfish. Journal of Virological Methods 2005, 123 (1), 1–7.

(26) Gregory, J. B.; Litaker, R. W.; Noble, R. T. Rapid One-Step Quantitative Reverse Transcriptase PCR Assay with Competitive Internal Positive Control for Detection of Enteroviruses in Environmental Samples. Applied and Environmental Microbiology 2006, 72 (6), 3960–3967.

(27) Lanciotti, R. S.; Kerst, A. J.; Nasci, R. S.; Godsey, M. S.; Mitchell, C. J.; Savage, H. M.; Komar, N.; Panella, N. A.; Allen, B. C.; Volpe, K. E.; Davis, B. S.; Roehrig, J. T. Rapid Detection of West Nile Virus from Human Clinical Specimens, Field-Collected Mosquitoes, and Avian Samples by a TaqMan Reverse Transcriptase-PCR Assay. Journal of Clinical Microbiology 2000, 38 (11), 4066–4071. 10.1128/jcm.38.11.4066-4071.2000.

(28) Boehm, A. B.; Wolfe, M. K.; Wigginton, K. R.; Bidwell, A.; White, B. J.; Hughes, B.; Duong; Chan-Herur, V.; Bischel, H. N.; Naughton, C. C. Human Viral Nucleic Acids Concentrations in Wastewater Solids from Central and Coastal California USA. Scientific Data 2023, 10, 396.

(29) Topol, A.; Wolfe, M.; White, B.; Wigginton, K.; Boehm, A. High Throughput Pre-Analytical Processing of Wastewater Settled Solids for SARS-CoV-2 RNA Analyses. protocols.io 2021. 10.17504/protocols.io.btyqnpvw.

(30) Topol, A.; Wolfe, M.; Wigginton, K.; White, B.; Boehm, A. High Throughput RNA Extraction and PCR Inhibitor Removal of Settled Solids for Wastewater Surveillance of SARS-CoV-2 RNA. protocols.io 2021. 10.17504/protocols.io.btyrnpv6.

(31) Huisman, J. S.; Scire, J.; Caduff, L.; Fernandez-Cassi, X.; Ganesanandamoorthy, P.; Kull, A.; Scheidegger, A.; Stachler, E.; Boehm, A. B.; Hughes, B.; Knudson, A.; Topol, A.; Wigginton, K. R.; Wolfe, M. K.; Kohn, T.; Ort, C.; Stadler, T.; Julian, T. R. Wastewater-Based Estimation of the Effective Reproductive Number of SARS-CoV-2. Environmental Health Perspectives 2022, 130 (5), 057011–1.

(32) Symonds, E. M.; Nguyen, K. H.; Harwood, V. J.; Breitbart, M. Pepper Mild Mottle Virus: A Plant Pathogen with a Greater Purpose in (Waste)Water Treatment Development and Public Health Management. Water Research 2018, 144, 1–12. 10.1016/j.watres.2018.06.066.

(33) Simpson, A.; Boehm, A. B. Effect of storage on concentrations of SARS-CoV-2 RNA in settled solids of wastewater treatment plants. Stanford Digital Repository. purl.stanford.edu/yn042kx5009.

(34) Borchardt, M. A.; Boehm, A. B.; Salit, M.; Spencer, S. K.; Wigginton, K. R.; Noble, R. T. The Environmental Microbiology Minimum Information (EMMI) Guidelines: QPCR and DPCR Quality and Reporting for Environmental Microbiology. Environ. Sci. Technol. 2021, 55 (15), 10210–10223. 10.1021/acs.est.1c01767.

(35) Kantor, R. S.; Nelson, K. L.; Greenwald, H. D.; Kennedy, L. C. Challenges in Measuring the Recovery of SARS-CoV-2 from Wastewater. Environ. Sci. Technol. 2021, 55 (6), 3514–3519. 10.1021/acs.est.0c08210.

(36) Fisman, D. Seasonality of Viral Infections: Mechanisms and Unknowns. Clinical Microbiology and Infection 2012, 18 (10), 946–954. 10.1111/j.1469-0691.2012.03968.x.

(37) Gholipour, S.; Ghalhari, M. R.; Nikaeen, M.; Rabbani, D.; Pakzad, P.; Miranzadeh, M. B. Occurrence of Viruses in Sewage Sludge: A Systematic Review. Science of The Total Environment 2022, 824, 153886. 10.1016/j.scitotenv.2022.153886.

(38) Bibby, K.; Peccia, J. Identification of Viral Pathogen Diversity in Sewage Sludge by Metagenome Analysis. Environ. Sci. Technol. 2013, 47 (4), 1945–1951. 10.1021/es305181x.

(39) Kittigul, L.; Pombubpa, K. Rotavirus Surveillance in Tap Water, Recycled Water, and Sewage Sludge in Thailand: A Longitudinal Study, 2007–2018. Food and Environmental Virology 2021, 13 (1), 53–63. 10.1007/s12560-020-09450-0.

(40) Silva-Sales, M.; Martínez-Puchol, S.; Gonzales-Gustavson, E.; Hundesa, A.; Gironès, R. High Prevalence of Rotavirus A in Raw Sewage Samples from Northeast Spain. Viruses 2020, 12 (3). 10.3390/v12030318.

(41) Zhou, N.; Lv, D.; Wang, S.; Lin, X.; Bi, Z.; Wang, H.; Wang, P.; Zhang, H.; Tao, Z.; Hou, P.; Song, Y.; Xu, A. Continuous Detection and Genetic Diversity of Human Rotavirus A in Sewage in Eastern China, 2013–2014. Virology Journal 2016, 13 (1), 153. 10.1186/s12985-016-0609-0.

(42) Barril, P. A.; Fumian, T. M.; Prez, V. E.; Gil, P. I.; Martínez, L. C.; Giordano, M. O.; Masachessi, G.; Isa, M. B.; Ferreyra, L. J.; Ré, V. E.; Miagostovich, M.; Pavan, J. V.; Nates, S. V. Rotavirus Seasonality in Urban Sewage from Argentina: Effect of Meteorological Variables on the Viral Load and the Genetic Diversity. Environmental Research 2015, 138, 409–415. 10.1016/j.envres.2015.03.004.

(43) Bosch, A.; Pinto, R. M.; Blanch, A. R.; Jofre, J. T. Detection of Human Rotavirus in Sewage through Two Concentration Procedures. Water Research 1988, 22 (3), 343–348. 10.1016/S0043-1354(88)90200-X.

(44) Hejkal, T. W.; Smith, E. M.; Gerba, C. P. Seasonal Occurrence of Rotavirus in Sewage. Applied and Environmental Microbiology 1984, 47 (3), 588–590. 10.1128/aem.47.3.588-590.1984.

(45) Rha, B.; Tate, J. E.; Payne, D. C.; Cortese, M. M.; Lopman, B. A.; Curns, A. T.; Parashar, U. D. Effectiveness and Impact of Rotavirus Vaccines in the United States – 2006–2012. Expert Review of Vaccines 2014, 13 (3), 365–376. 10.1586/14760584.2014.877846.

(46) Anderson, E. J. Rotavirus Vaccines: Viral Shedding and Risk of Transmission. The Lancet Infectious Diseases 2008, 8 (10), 642–649. 10.1016/S1473-3099(08)70231-7.

(47) Eftim, S. E.; Hong, T.; Soller, J.; Boehm, A.; Warren, I.; Ichida, A.; Nappier, S. P. Occurrence of Norovirus in Raw Sewage – A Systematic Literature Review and Meta-Analysis. Water Research 2017, 111 (Supplement C), 366–374. 10.1016/j.watres.2017.01.017.

(48) Betancourt, W. Q.; Shulman, L. M. Polioviruses and Other Enteroviruses. In Water and Sanitation for the 21st Century: Health and Microbiological Aspects of Excreta and Wastewater Management (Global Water Pathogen Project; Rose, J. B., Jimenez-Cisneros, B., Eds.; Meschke, J. S., Girones, R., Series Eds.; Part 3: Specific Excreted Pathogens: Environmental and Epidemiology Aspects - Section 1: Viruses; Michigan State University, East Lansing, MI, 2017.

(49) Brinkman, N. E.; Shay, F. G.; Keely, S. P. Retrospective Surveillance of Wastewater To Examine Seasonal Dynamics of Enterovirus Infections. mSphere 2017, 2 (3), e00099–17. 10.1128/mSphere.00099-17.

(50) Monpoeho, S.; Dehée, A.; Mignotte, B.; Schwartzbrod, L.; Marechal, V.; Nicolas, J.-C.; Billaudel, S.; Férré, V. Quantification of Enterovirus RNA in Sludge Samples Using Single Tube Real-Time RT-PCR. BioTechniques 2000, 29 (1), 88–93. 10.2144/00291st03.

(51) Gerba, C. P.; Goyal, S. M.; Hurst, C. J.; Labelle, R. L. Type and Strain Dependence of Enterovirus Adsorption to Activated Sludge, Soils and Estuarine Sediments. Water Research 1980, 14 (9), 1197–1198. 10.1016/0043-1354(80)90176-1.

(52) Fong, T.-T.; Phanikumar, M. S.; Xagoraraki, I.; Rose, J. B. Quantitative Detection of Human Adenoviruses in Wastewater and Combined Sewer Overflows Influencing a Michigan River. Applied and Environmental Microbiology 2010, 76 (3), 715–723. 10.1128/AEM.01316-09.

(53) Katayama, H.; Haramoto, E.; Oguma, K.; Yamashita, H.; Tajima, A.; Nakajima, H.; Ohgaki, S. One-Year Monthly Quantitative Survey of Noroviruses, Enteroviruses, and Adenoviruses in Wastewater Collected from Six Plants in Japan. Water Research 2008, 42 (6), 1441–1448. 10.1016/j.watres.2007.10.029.

(54) Jiang, S. C. Human Adenoviruses in Water: Occurrence and Health Implications: A Critical Review. Environ. Sci. Technol. 2006, 40 (23), 7132–7140. 10.1021/es060892o.

(55) Martin, N. A.; Gonzalez, G.; Reynolds, L. J.; Bennett, C.; Campbell, C.; Nolan, T. M.; Byrne, A.; Fennema, S.; Holohan, N.; Kuntamukkula, S. R.; Sarwar, N.; Sala-Comorera, L.; Dean, J.; Urtasun-Elizari, J. M.; Hare, D.; Liddy, E.; Joyce, E.; O’Sullivan, J. J.; Cuddihy, J. M.; McIntyre, A. M.; Robinson, E. P.; Dahly, D.; Fletcher, N. F.; Cotter, S.; Fitzpatrick, E.; Carr, M. J.; De Gascun, C. F.; Meijer, W. G. Adeno-Associated Virus 2 and Human Adenovirus F41 in Wastewater during Outbreak of Severe Acute Hepatitis in Children, Ireland. Emerg Infect Dis 2023, 29 (4), 751–760. 10.3201/eid2904.221878.

(56) Reyne, M. I.; Allen, D. M.; Levickas, A.; Allingham, P.; Lock, J.; Fitzgerald, A.; McSparron, C.; Nejad, B. F.; McKinley, J.; Lee, A.; Bell, S. H.; Quick, J.; Houldcroft, C. J.; Bamford, C. G. G.; Gilpin, D. F.; McGrath, J. W. Detection of Human Adenovirus F41 in Wastewater and Its Relationship to Clinical Cases of Acute Hepatitis of Unknown Aetiology. Science of The Total Environment 2023, 857, 159579. 10.1016/j.scitotenv.2022.159579.

(57) Brooks, G.; Butel, J.; Morse, S. Medical Microbiology, 22nd ed.; McGraw-Hill: San Francisco, CA, 2001.

(58) Takuissu, G. R.; Kenmoe, S.; Ebogo-Belobo, J. T.; Kengne-Ndé, C.; Mbaga, D. S.; Bowo-Ngandji, A.; Ndzie Ondigui, J. L.; Kenfack-Momo, R.; Tchatchouang, S.; Kenfack-Zanguim, J.; Lontuo Fogang, R.; Zeuko’o Menkem, E.; Kame-Ngasse, G. I.; Magoudjou-Pekam, J. N.; Veneri, C.; Mancini, P.; Bonanno Ferraro, G.; Iaconelli, M.; Orlandi, L.; Del Giudice, C.; Suffredini, E.; La Rosa, G. Occurrence of Hepatitis A Virus in Water Matrices: A Systematic Review and Meta-Analysis. Int J Environ Res Public Health 2023, 20 (2). 10.3390/ijerph20021054.

(59) Hellmér, M.; Paxéus, N.; Magnius, L.; Enache, L.; Arnholm, B.; Johansson, A.; Bergström, T.; Norder, H. Detection of Pathogenic Viruses in Sewage Provided Early Warnings of Hepatitis A Virus and Norovirus Outbreaks. Applied and Environmental Microbiology 2014, 80 (21), 6771–6781. 10.1128/AEM.01981-14.

(60) Arraj, A.; Bohatier, J.; Laveran, H.; Traore, O. Comparison of Bacteriophage and Enteric Virus Removal in Pilot Scale Activated Sludge Plants. Journal of Applied Microbiology 2005, 98 (2), 516–524. 10.1111/j.1365-2672.2004.02485.x.

(61) Tisza, M.; Cregeen, S. J.; Avadhanula, V.; Zhang, P.; Ayvaz, T.; Feliz, K.; Hoffman, K. L.; Clark, J. R.; Terwilliger, A.; Ross, M. C.; Cormier, J.; Henke, D.; Troisi, C.; Wu, F.; Rios, J.; Deegan, J.; Hansen, B.; Balliew, J.; Gitter, A.; Zhang, K.; Li, R.; Bauer, C. X.; Mena, K. D.; Piedra, P. A.; Petrosino, J. F.; Boerwinkle, E.; Maresso, A. W. Comprehensive Wastewater Sequencing Reveals Community and Variant Dynamics of the Collective Human Virome. medRxiv 2023. 10.1101/2023.05.03.23289441.

(62) Erster, O.; Bar-Or, I.; Levy, V.; Shatzman-Steuerman, R.; Sofer, D.; Weiss, L.; Vasserman, R.; Fratty, I. S.; Kestin, K.; Elul, M.; Levi, N.; Alkrenawi, R.; Mendelson, E.; Mandelboim, M.; Weil, M. Monitoring of Enterovirus D68 Outbreak in Israel by a Parallel Clinical and Wastewater Based Surveillance. Viruses 2022, 14 (5). 10.3390/v14051010.

(63) Tedcastle, A.; Wilton, T.; Pegg, E.; Klapsa, D.; Bujaki, E.; Mate, R.; Fritzsche, M.; Majumdar, M.; Martin, J. Detection of Enterovirus D68 in Wastewater Samples from the UK between July and November 2021. Viruses 2022, 14 (1). 10.3390/v14010143.

(64) Vila, T.; Sultan, A. S.; Montelongo-Jauregui, D.; Jabra-Rizk, M. A. Candida Auris: A Fungus with Identity Crisis. Pathogens and Disease 2020, 78 (4), ftaa034. 10.1093/femspd/ftaa034.

(65) Osei Sekyere, J. Candida Auris: A Systematic Review and Meta-Analysis of Current Updates on an Emerging Multidrug-Resistant Pathogen. Microbiologyopen 2018, 7 (4), e00578. 10.1002/mbo3.578.

(66) Lee, W. L.; Gu, X.; Armas, F.; Leifels, M.; Wu, F.; Chandra, F.; Chua, F. J. D.; Syenina, A.; Chen, H.; Cheng, D.; Ooi, E. E.; Wuertz, S.; Alm, E. J.; Thompson, J. Monitoring Human Arboviral Diseases through Wastewater Surveillance: Challenges, Progress and Future Opportunities. Water Research 2022, 223, 118904. 10.1016/j.watres.2022.118904.

