## Supporting information for "Two years of longitudinal measurements of human adenovirus group F, norovirus GI and GII, rotavirus, enterovirus, enterovirus D68, hepatitis A virus, *Candida auris*, and West Nile virus nucleic-acids in wastewater solids: A retrospective study at two wastewater treatment plants"

**Additional details related to the EMMI guidelines.** Thirty-six samples were selected at random for this analysis; this represents 8% of the samples processed in the study. As described in the methods section, each sample was run as template in three different PCR reactions; 1 for PMMoV, 1 for rotavirus, SARS-CoV-2, WNV, HuNoV GII, and HAdV, and 1 for EVD68, HAV, EV, HuNoV GI, and C. auris. The average (standard deviation) number of partitions (droplets) for each of the three reactions (across the 10 replicates) was 164164 (38851) for the reaction for PMMoV, 174321 (20126) for the reaction for rotavirus, SARS-CoV-2, WNV, HuNoV GII, and HAdV, and 185497 (24410) for the reaction for EVD68, HAV, EV, HuNoV GI, and C. auris. The volume of the partitions, as reported by the machine vendor is 0.00085 µL. The mean and standard deviation of copies per partition for each target is shown in Table S2.

Table S1. Parameters used in the development of new primers and probes using Primer3Plus (https://primer3plus.com/, accessed 7/16/23).

- Product size ranges: 60-275
- Primer size: min 15, opt 20, max 36
- Primer melting temperature: min 50°C, optimal 60°C, max 65°C - GC% content: min 40%, optimal 50%, high 60%
- concentration of divalent cations = 3.8 mM
- concentration of dNTPs needs to be 0.8 mM
- Internal Oligo: size min 15, optimal 20, max 30
- Internal Oligo: Melting temp min 62°C, optimal 63°C, max 70°C
- Internal Oligo: GC% min 30%, optimal 50%, max 80%

Table S2. For each target measured in this study, the mean and standard deviation (sd) of the total number of copies of target per partition. Num is the number of samples out of a random 36 included in this analysis that had detectable target in them and thus contributed to the calculated mean and standard deviation. A value of 0 indicates that of the random 36 samples selected, none of them had the target present in them. Abbreviations for the targets are provided in the main text except "Rota" is rotavirus and SC2 is the N gene of SARS-CoV-2.

| Target | EVD68 | HAV | EV | HuNoV GI | C. auris | Rota | SC2 | WNV | HuNoV GII | HAdV | PMMoV |
|---|---|---|---|---|---|---|---|---|---|---|---|
| mean | $1.04x10^{-3}$ | $9.91x10^{-5}$ | $1.02x10^{-3}$ | $8.93x10^{-4}$ | 0 | $1.20x10^{-4}$ | $1.37x10^{-3}$ | 0 | $1.12x10^{-2}$ | $5.03x10^{-2}$ | 0.15 |
| sd | $1.18x10^{-3}$ | $1.59x10^{-4}$ | $3.65x10^{-4}$ | $7.94x10^{-4}$ | 0 | $1.14x10^{-4}$ | $1.22x10^{-3}$ | 0 | $8.57x10^{-3}$ | $3.58x10^{-2}$ | 0.069 |
| num | 19 | 15 | 36 | 36 | 0 | 23 | 36 | 0 | 36 | 36 | 36 |

Figure S1. EMMI checklist.

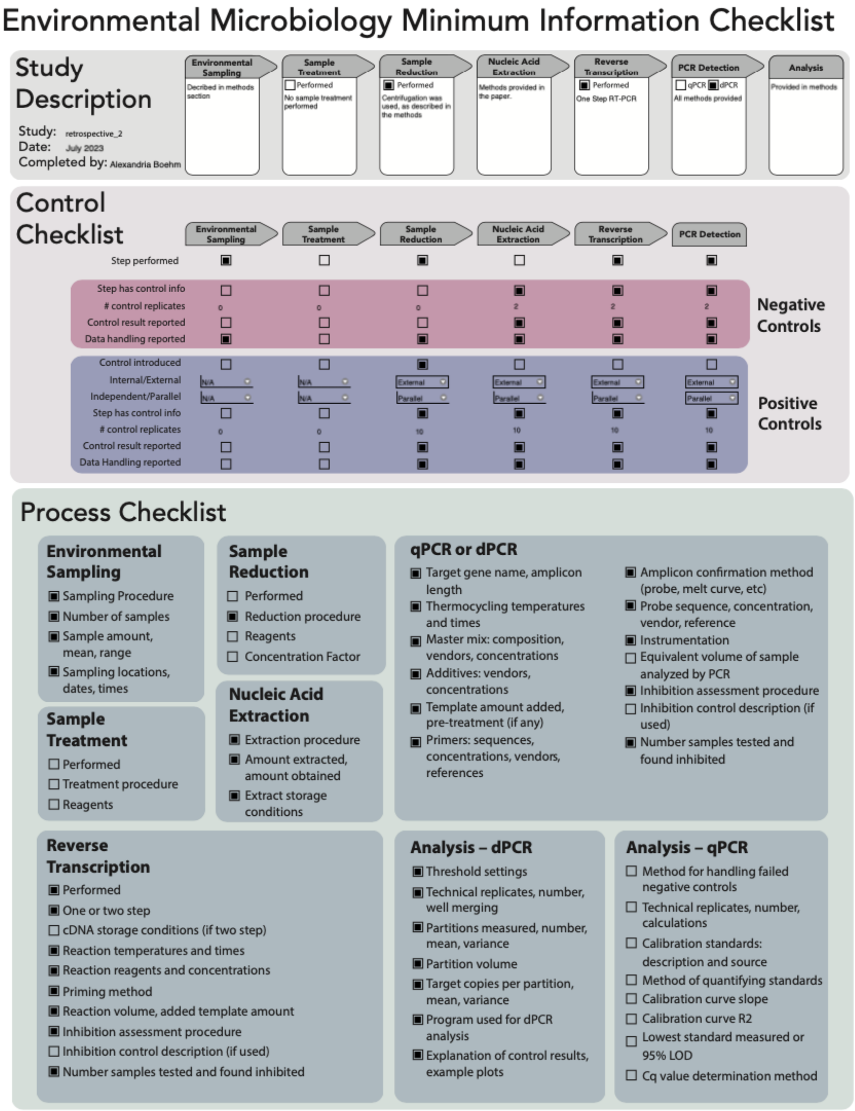

### Environmental Microbiology Minimum Information Checklist

#### Study Description

Study: retrospective_2
Date: July 2023
Completed by: Alexandria Boehm

| Environmental Sampling | Sample Treatment | Sample Reduction | Nucleic Acid Extraction | Reverse Transcription | PCR Detection | Analysis |
|---|---|---|---|---|---|---|
| Decribed in methods section | ☐ Performed<br>No sample treatment performed | ■ Performed<br>Centrifugation was used, as described in the methods | Methods provided in the paper. | ■ Performed<br>One Step RT-PCR | ☐ qPCR ■ dPCR<br>All methods provided | Provided in methods |

#### Control Checklist

| | Environmental Sampling | Sample Treatment | Sample Reduction | Nucleic Acid Extraction | Reverse Transcription | PCR Detection |
|---|---|---|---|---|---|---|
| Step performed | ■ | ☐ | ■ | ☐ | ■ | ■ |
| **Negative Controls** | | | | | | |
| Step has control info | ☐ | ☐ | ☐ | ■ | ■ | ■ |
| # control replicates | 0 | 0 | 0 | 2 | 2 | 2 |
| Control result reported | ☐ | ☐ | ☐ | ■ | ■ | ■ |
| Data handling reported | ■ | ☐ | ■ | ■ | ■ | ■ |
| **Positive Controls** | | | | | | |
| Control introduced | ☐ | ☐ | ■ | ☐ | ☐ | ☐ |
| Internal/External | N/A | N/A | External | External | External | External |
| Independent/Parallel | N/A | N/A | Parallel | Parallel | Parallel | Parallel |
| Step has control info | ☐ | ☐ | ■ | ■ | ■ | ■ |
| # control replicates | 0 | 0 | 10 | 10 | 10 | 10 |
| Control result reported | ☐ | ☐ | ■ | ■ | ■ | ■ |
| Data Handling reported | ☐ | ☐ | ■ | ■ | ■ | ■ |

#### Process Checklist

##### Environmental Sampling
- ■ Sampling Procedure
- ■ Number of samples
- ■ Sample amount, mean, range
- ■ Sampling locations, dates, times

##### Sample Treatment
- ☐ Performed
- ☐ Treatment procedure
- ☐ Reagents

##### Sample Reduction
- ☐ Performed
- ■ Reduction procedure
- ☐ Reagents
- ☐ Concentration Factor

##### Nucleic Acid Extraction
- ■ Extraction procedure
- ■ Amount extracted, amount obtained
- ■ Extract storage conditions

##### qPCR or dPCR
- ■ Target gene name, amplicon length
- ■ Thermocycling temperatures and times
- ■ Master mix: composition, vendors, concentrations
- ■ Additives: vendors, concentrations
- ■ Template amount added, pre-treatment (if any)
- ■ Primers: sequences, concentrations, vendors, references
- ■ Amplicon confirmation method (probe, melt curve, etc)
- ■ Probe sequence, concentration, vendor, reference
- ■ Instrumentation
- ☐ Equivalent volume of sample analyzed by PCR
- ■ Inhibition assessment procedure
- ☐ Inhibition control description (if used)
- ■ Number samples tested and found inhibited

##### Reverse Transcription
- ■ Performed
- ■ One or two step
- ☐ cDNA storage conditions (if two step)
- ■ Reaction temperatures and times
- ■ Reaction reagents and concentrations
- ■ Priming method
- ■ Reaction volume, added template amount
- ■ Inhibition assessment procedure
- ☐ Inhibition control description (if used)
- ■ Number samples tested and found inhibited

##### Analysis – dPCR
- ■ Threshold settings
- ■ Technical replicates, number, well merging
- ■ Partitions measured, number, mean, variance
- ■ Partition volume
- ■ Target copies per partition, mean, variance
- ■ Program used for dPCR analysis
- ■ Explanation of control results, example plots

##### Analysis – qPCR
- ☐ Method for handling failed negative controls
- ☐ Technical replicates, number, calculations
- ☐ Calibration standards: description and source
- ☐ Method of quantifying standards
- ☐ Calibration curve slope
- ☐ Calibration curve R2
- ☐ Lowest standard measured or 95% LOD
- ☐ Cq value determination method

Figure S2. Concentrations of HuNoV G1 and GII, rotavirus, HAdV, EV, HAV, EVD68 and *C. auris* in wastewater solids at OSP and SJ normalized by concentrations of PMMoV. Error bars show standard deviation of the ratio propagated by assuming the errors are symmetric and described by the larger of the lower or error bar from the original measurements. The black line shows a 5-adjacent sample trimmed average. A gray symbol at the top of the plot indicates a value that is higher than the y-axis scale.

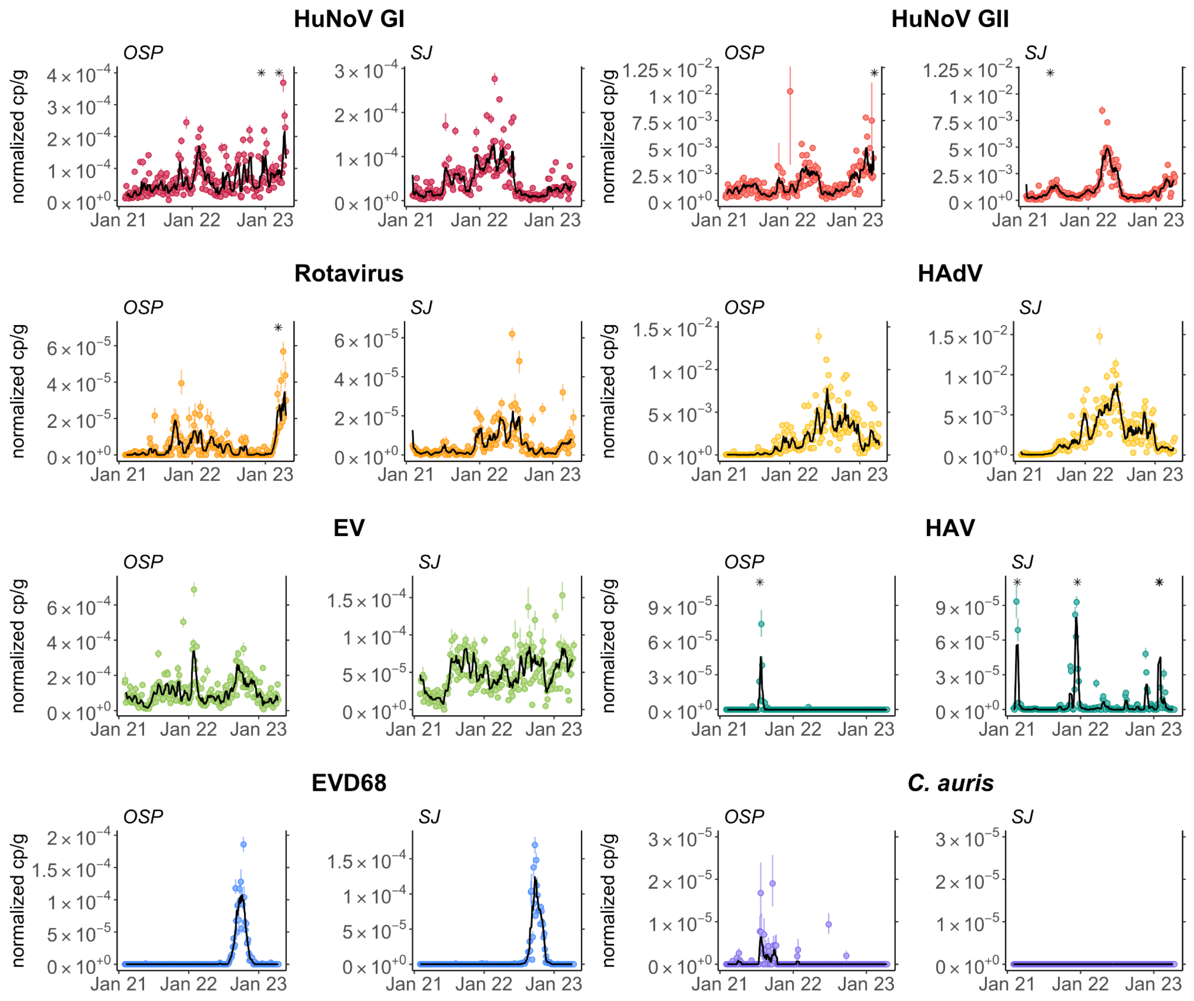
